# COVID-19 Myocardial Pathology Evaluated Through scrEening Cardiac Magnetic Resonance (COMPETE CMR)

**DOI:** 10.1101/2020.08.31.20185140

**Authors:** Daniel E. Clark, Amar Parikh, Jeffrey M. Dendy, Alex B. Diamond, Kristen George-Durrett, Frank A. Fish, Warne Fitch, Sean G. Hughes, Jonathan H. Soslow

**Author notes:** Co-senior authors listed in alphabetical order (Sean G. Hughes, Jonathan H. Soslow). **Address for correspondence:** Daniel E. Clark, MD, MPH, 2220 Pierce Avenue, 383 Preston Research Building, Nashville, TN 37237, @DanClarkMD. **Tweet:** Cardiac sequelae among athletes with asymptomatic or mild COVID-19 only 9% by #whyCMR, but may occur without symptoms and be missed by #echofirst.

## Abstract

**Background:** Myocarditis is a leading cause of sudden cardiac death among competitive athletes and may occur without antecedent symptoms. COVID-19-associated myocarditis has been well-described, but the prevalence of myocardial inflammation and fibrosis in young athletes after COVID-19 infection is unknown.

**Objectives:** This study sought to evaluate the prevalence and extent of cardiovascular involvement in collegiate athletes that had recently recovered from COVID-19.

**Methods:** We conducted a retrospective cohort analysis of collegiate varsity athletes with prior COVID-19 infection, all of whom underwent cardiac magnetic resonance (CMR) prior to resumption of competitive sports in August 2020.

**Results:** Twenty-two collegiate athletes with prior COVID-19 infection underwent CMR. The median time from SARS-CoV-2 infection to CMR was 52 days. The mean age was 20.2 years. Athletes represented 8 different varsity sports. This cohort was compared to 22 healthy controls and 22 tactical athlete controls. Most athletes experienced mild illness (N=17, 77%), while the remainder (23%) were asymptomatic. No athletes had abnormal troponin I, electrocardiograms, or LVEF < 50% on echocardiography. Late gadolinium enhancement was found in 9% of collegiate athletes and one athlete (5%) met formal criteria for myocarditis.

**Conclusions:** Our study suggests that the prevalence of myocardial inflammation or fibrosis after an asymptomatic or mild course of ambulatory COVID-19 among competitive athletes is modest (9%), but would be missed by ECG, Ti, and strain echocardiography. Future investigation is necessary to further phenotype cardiovascular manifestations of COVID-19 in order to better counsel athletes on return to sports participation.

**Condensed Abstract:** COVID-19-associated myocarditis has been well-described, but the prevalence of myocardial inflammation and fibrosis in athletes after COVID-19 is unknown. We conducted a retrospective cohort analysis of 22 collegiate athletes with prior COVID-19 infection who underwent electrocardiography, troponin I, echocardiography with strain, and CMR. The median time from SARS-CoV-2 infection to CMR was 52 days. All athletes experienced mild illness or were asymptomatic. Late gadolinium enhancement was found in 9%. This suggests the prevalence of myocardial inflammation or fibrosis after an asymptomatic or mild course of COVID-19 among competitive athletes is modest, but would be missed without CMR screening.

## INTRODUCTION

Over 25 million people worldwide have now tested positive for coronavirus disease-19 (COVID-19), and over 843,000 have died of severe acute respiratory syndrome coronavirus-2 (SARS-CoV-2).(1) Though respiratory symptoms predominate in the early course of disease, cardiovascular complications are increasingly recognized as a cause of morbidity and mortality later in the disease process. Pre-existing cardiovascular disease and traditional cardiac risk factors have been shown to increase the risk of cardiac complications of COVID-19 infection, but even healthy and asymptomatic COVID-19 survivors have suffered cardiac complications.(2) Echocardiography has revealed a high prevalence (8%) of myocardial infarction, myocarditis, or Takotsubo cardiomyopathy among hospitalized patients with presumed or confirmed COVID-19 infection.(3) Most recently, in a large, predominantly ambulatory cohort, cardiac magnet resonance (CMR) revealed myopericardial inflammation or scarring in 78 of 100 patients after recovery from COVID-19 infection.(4)

As school and athletics resume on many college campuses this fall, the safe return of student-athletes to sport is a major public health concern. Although competitive athletes may be less likely to suffer morbidity during acute COVID-19 infection than the general population, they may be at heightened risk for malignant arrhythmia during strenuous exertion from sequelae of COVID-19 associated myocarditis.(5) The prevalence of persistent myocardial inflammation and fibrosis in young athletes after COVID-19 infection is not known. Meanwhile, myocarditis represents the third leading cause of SCD among US competitive athletes, and may occur without any antecedent symptoms.(6,7) Athletic departments and academic centers are devising return-to-competition guidelines without clear data on the prevalence of cardiovascular abnormalities after COVID-19. We sought to use comprehensive cardiac magnetic resonance (CMR) to assess the prevalence and extent of cardiovascular sequelae in collegiate athletes that had recently recovered at home from COVID-19 infection.

## METHODS

### Study design and patient population

We conducted a retrospective cohort analysis of varsity athletes from a single National Collegiate Athletic Association (NCAA) Division 1 Power 5 institution referred for CMR after COVID-19 infection. Since August 2020 varsity athletes at this institution were screened for SARS-CoV-2 infection upon return to campus and twice weekly (real-time polymerase chain reaction nasal swab), regardless of symptomatology. The frequency of screening is anticipated to increase with return of the Southeastern Conference fall athletic season, as all athletes must have a negative SARS-CoV-2 test 72 hours prior to competition. In all COVID-19+ athletes, the protocol necessitates an electrocardiogram read by a Sports Cardiologist, troponin I, echocardiogram with strain imaging, and contrasted CMR. Only athletes over 18 years of age were included in this analysis. Clinical demographics, laboratory, electrocardiographic, and CMR results were stored in the REDCap electronic platform.(8)

Healthy controls were used from a cohort of healthy adult subjects over 18 years old who had previously consented for non-contrasted CMR imaging, including parametric mapping; these controls were a subset of the subjects used to derive our laboratory’s normal control values. Additional athletic controls were selected from a cohort of tactical athletes referred to our center over the past 12 months for cardiac symptomatology found to have normal cardiac function without pathology. As most controls were non-contrast or did not have available hematocrit, ECV values were compared to published reference ranges. The mid-septal ECV from “The International T1 Multicenter Cardiovascular Magnetic Resonance Study” was used for healthy control group comparison (Group 1, < 30 years old, N=27 for 1.5-T).(9) Given that cardiomyocyte hypertrophy among athletes is known to result in lower ECV than healthy controls, literature normative values for ECV among athletes was used for comparison for the athletic control group (N=30, mean age 28 years).(10) No controls (healthy or tactical athletic) underwent routine testing with ECG, cardiac biomarkers, or echocardiography. Tactical athletes did not undergo T_2_ mapping.

Our laboratory’s normal values for parametric mapping are derived from a cohort of 54 healthy controls of varying age (range 7-56 years old) and gender (N=29 male) prospectively enrolled and then broken down into the appropriate gender and age (>18 y/o and <18 y/o) category for comparison. The normal ranges were derived from the mean and standard deviation by setting 2 standard deviations above the mean as the upper range of normal. For this manuscript, we considered presence of any LGE abnormal. LVEF 50-55% was not considered abnormal given the possibility of borderline to mildly depressed function in patients with athlete’s heart.(11)

### CMR protocol

A comprehensive CMR with contrast was performed on a 1.5 Tesla Siemens Avanto Fit (Siemens Healthcare Sector, Erlangen, Germany). The CMR protocol consisted of cine CMR balanced stead-state free precession imaging to calculate left and right ventricular volumes, left ventricular ejection fraction (LVEF), and myocardial mass. Intravenous gadolinium contrast (gadobutrol, Gadavist®, Bayer Healthcare Pharmaceuticals, Wayne, NJ, USA at a dose of 0.15 mmol/kg) was administered through a peripheral intravenous line. Late gadolinium enhancement (LGE) was performed using segmented inversion recovery (optimized inversion time to null myocardium) and single shot phase sensitive inversion recovery (inversion time of 300ms) imaging in standard long-axis planes and a short-axis stack. Native T_1_ mapping, T_2_ mapping, and post-contrast (15 minutes after contrast administration) T_1_ mapping was performed. T_1_ mapping was performed using a modified Look-Locker inversion recovery (MOLLI) sequence acquired as using 5(3s)3 protocol before contrast and 4(1)3(1)2 protocol after contrast (see supplementary material for more details).

### CMR post-processing

CMR post-processing was performed blinded to clinical data. Ventricular volumes and function were calculated using Medis QMass (MedisSuite 2.1, Medis, Leiden, The Netherlands). The presence or absence of LGE, as well as location using the standard 17-segment model,(12) was qualitatively assessed by two cardiologists with over 10 years of experience reading CMR. If there was disagreement, a third cardiologist assessed for LGE.

T_1_ maps were obtained prior to and after contrast administration as described by Messroghli et al.(13) Regions of interest (ROIs) were manually drawn on T_1_, T_2_, and ECV maps within the LV mesocardium, carefully avoiding partial volume averaging with blood-pool or epicardial fat and artifact. As per our lab protocol, ROIs were placed in the septum and free wall of the basal and mid short-axis slices, as well as in areas with focal abnormalities identified after visual inspection. Areas of LGE determined not to be due to myocardial infarction were included in the analysis, as these were felt to be the most focal areas in a continuum of diffuse ECM expansion.(14)

### Statistical analysis

Categorical variables were compared using the chi squared test and presented as frequency and percentage. Continuous variables were compared using the Wilcoxon rank sum and presented as median and interquartile range (IQR). For statistical comparisons with reported reference ranges (ECV), a Student t-test was used to compare the mean, standard deviation, and sample size of published values to those of our patient population. Statistical analysis was performed using STATA, version 15 (StatCorp LLC, College Station, TX) software. All tests were 2-sided and a p-value < 0.05 was considered significant. The study was approved by the institutional review board at Vanderbilt University Medical Center.

## RESULTS

Twenty-two competitive athletes with COVID-19 (COVID-19+ athletes) and 44 controls (22 healthy controls and 22 tactical athletes) were included. Tactical athletic controls were more likely to be older age, male gender, and have larger body surface areas (BSAs) compared to COVID-19+ athletes. The COVID-19+ athletes represented 8 different collegiate sports: lacrosse (1), swimming (1), tennis (2), football (2), baseball (2), basketball (3), golf (3), and soccer (8). The COVID-19+ athlete group was 59% female and 25% non-white, with a median age of 20 years. The majority of COVID-19+ athletes experienced mild illness (N=17, 77%), while 23% were asymptomatic and diagnosed by routine screening. The median time from SARS-CoV-2 infection to CMR was 52 days.

### ECG, cardiac biomarker, and echocardiography findings

Twenty (91%) of COVID-19+ athletes had a screening ECG and 18 (82%) had a troponin I collected and all were within normal limits (troponin I < 99% for age). All additional cardiac biomarkers (brain natriuretic peptide in 3 and high-sensitivity c-reactive protein in 2) were within the normal range. Echocardiography was performed in 21 COVID-19+ athletes and strain imaging was completed in 16. The median LVEF was 59% and the median global longitudinal strain was within the normal range at −18.2% (**Table 1**).

**Table 1.**
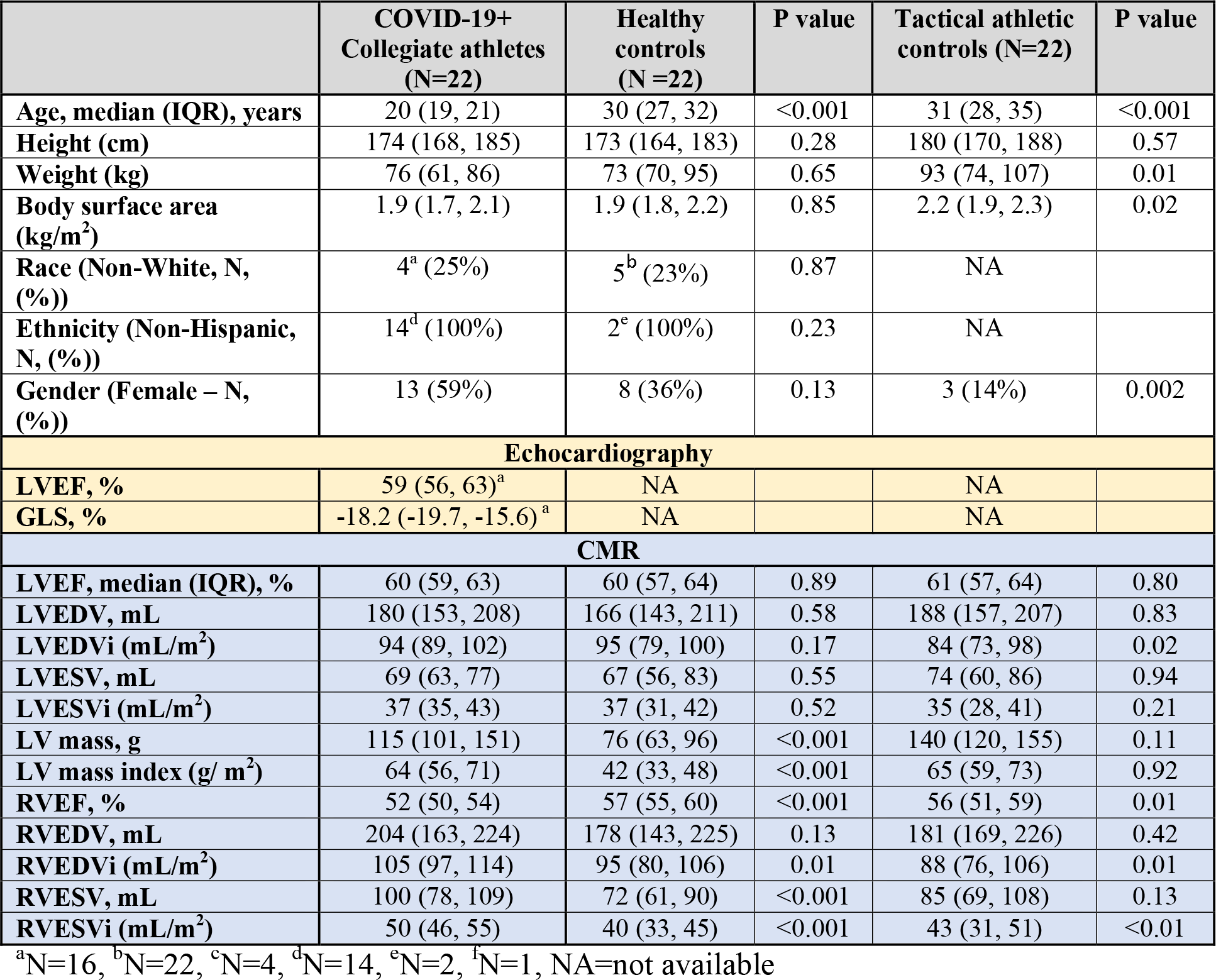
Baseline characteristics.

|  | **COVID-19+ Collegiate athletes (N=22)** | **Healthy controls (N =22)** | **P value** | **Tactical athletic controls (N=22)** | **P value** |
| --- | --- | --- | --- | --- | --- |
| **Age, median (IQR), years** | 20 (19, 21) | 30 (27, 32) | <0.001 | 31 (28, 35) | <0.001 |
| **Height (cm)** | 174 (168, 185) | 173 (164, 183) | 0.28 | 180 (170, 188) | 0.57 |
| **Weight (kg)** | 76 (61, 86) | 73 (70, 95) | 0.65 | 93 (74, 107) | 0.01 |
| **Body surface area (kg/m<sup>2</sup>)** | 1.9 (1.7, 2.1) | 1.9 (1.8, 2.2) | 0.85 | 2.2 (1.9, 2.3) | 0.02 |
| **Race (Non-White, N, (%))** | 4<sup>a</sup> (25%) | 5<sup>b</sup> (23%) | 0.87 | NA |  |
| **Ethnicity (Non-Hispanic, N, (%))** | 14<sup>d</sup> (100%) | 2<sup>e</sup> (100%) | 0.23 | NA |  |
| **Gender (Female – N, (%))** | 13 (59%) | 8 (36%) | 0.13 | 3 (14%) | 0.002 |
| **Echocardiography** |  |  |  |  |  |
| **LVEF, %** | 59 (56, 63)<sup>a</sup> | NA |  | NA |  |
| **GLS, %** | -18.2 (-19.7, -15.6)<sup>a</sup> | NA |  | NA |  |
| **CMR** |  |  |  |  |  |
| **LVEF, median (IQR), %** | 60 (59, 63) | 60 (57, 64) | 0.89 | 61 (57, 64) | 0.80 |
| **LVEDV, mL** | 180 (153, 208) | 166 (143, 211) | 0.58 | 188 (157, 207) | 0.83 |
| **LVEDVi (mL/m<sup>2</sup>)** | 94 (89, 102) | 95 (79, 100) | 0.17 | 84 (73, 98) | 0.02 |
| **LVESV, mL** | 69 (63, 77) | 67 (56, 83) | 0.55 | 74 (60, 86) | 0.94 |
| **LVESVi (mL/m<sup>2</sup>)** | 37 (35, 43) | 37 (31, 42) | 0.52 | 35 (28, 41) | 0.21 |
| **LV mass, g** | 115 (101, 151) | 76 (63, 96) | <0.001 | 140 (120, 155) | 0.11 |
| **LV mass index (g/ m<sup>2</sup>)** | 64 (56, 71) | 42 (33, 48) | <0.001 | 65 (59, 73) | 0.92 |
| **RVEF, %** | 52 (50, 54) | 57 (55, 60) | <0.001 | 56 (51, 59) | 0.01 |
| **RVEDV, mL** | 204 (163, 224) | 178 (143, 225) | 0.13 | 181 (169, 226) | 0.42 |
| **RVEDVi (mL/m<sup>2</sup>)** | 105 (97, 114) | 95 (80, 106) | 0.01 | 88 (76, 106) | 0.01 |
| **RVESV, mL** | 100 (78, 109) | 72 (61, 90) | <0.001 | 85 (69, 108) | 0.13 |
| **RVESVi (mL/m<sup>2</sup>)** | 50 (46, 55) | 40 (33, 45) | <0.001 | 43 (31, 51) | <0.01 |

<sup>a</sup>N=16, <sup>b</sup>N=22, <sup>c</sup>N=4, <sup>d</sup>N=14, <sup>e</sup>N=2, <sup>f</sup>N=1, NA=not available

### CMR volumetric and functional comparisons

Cardiac volumes and function were compared among groups (**Table 1**). The left ventricular ejection fraction (LVEF) and volumetrics were normal and similar across groups, except for reduced LV end diastolic volume index (LVEDVi) among tactical athletic controls (84 ml/m^2^) compared to healthy controls (95 ml/m^2^) and COVID-19+ athletes (94 ml/m^2^). Healthy controls had a significantly lower LV mass index (42 g/m^2^) compared to COVID-19+ athletes (64 g/m^2^) and tactical athletic controls (65 g/m^2^). The right ventricular ejection fraction was significantly lower in COVID-19+ athletes (52%) compared to healthy controls (57%) and tactical athletic controls (56%). COVID-19+ athletes also had increased RV volumes relative to healthy controls and tactical athletes (**Table 1**).

### CMR parametric mapping and scar imaging

There was no difference in native T_1_ mapping among groups at any of the four ROIs (**Table 2**). T_2_ times were elevated in COVID-19+ patients compared to healthy controls (46.4 vs. 44.6 ms). Mid septal ECV was higher among COVID-19+ athletes (25.3, SD ± 2.6) as compared to reference values for both athletic controls (22.5, SD ± 3) and healthy controls (24.0, SD ± 3), though the latter did not reach statistical significance.

**Table 2.** CMR parametric mapping and LGE comparison.

|  | **COVID-19+ Collegiate athletes (N=22)** | **Healthy controls (N=22)** | **P value** | **Tactical athletic controls (N=22)** | **P value** |
| --- | --- | --- | --- | --- | --- |
| **T<sub>1</sub> (global), median (IQR), ms** |  |  |  |  |  |
| Basal septum | 995 (969, 1006) | 986 (971, 999) | 0.31 | 990 (964, 1024) | 0.89 |
| Basal lateral | 971 (951, 984) | 958 (945, 983) | 0.53 | 975 (962, 1011) | 0.17 |
| Mid septum | 982 (973, 997) | 978 (963, 998) | 0.62 | 989 (963, 1008) | 0.22 |
| Mid lateral | 980 (947, 988) | 965 (946, 975) | 0.20 | 968 (944, 1000) | 0.93 |
| **T<sub>2</sub> (global), ms** |  |  |  |  |  |
| Basal septum | 44.3 (42.3, 46.1) | 42.4 (41.5, 43.3) | 0.009 | * |  |
| Basal lateral | 45.4 (43.2, 46.6) | 44.0 (43.0, 44.6) | 0.034 | * |  |
| Mid septum | 46.4 (45.2, 48.2) | 44.6 (43.2, 45.4) | 0.004 | * |  |
| Mid lateral | 47.0 (45.2, 48.2) | 44.0 (42.6, 45.4) | 0.003 | * |  |
| **ECV (global), %** |  |  |  |  |  |
| Basal septum | 24.0 (22.7, 25.9) |  |  |  |  |
| Basal lateral | 21.6 (20.6, 23.8) |  |  |  |  |
| Mid septum | 25.4 (22.7, 27.3)<br>25.3 ± 2.6 (SD) | 24 (SD ± 3)<sup>b</sup> | 0.11 | 22.5 ± 2.6 (SD)<sup>c</sup> | <0.001<sup>¶</sup> |
| Mid lateral | 23.3 (21.4, 25.0) |  |  |  |  |
| **LGE (Any), N (%)** | 2 (9 %) | 0 |  | 0 |  |
| Myocardial | 2 (9 %) | 0 |  | 0 |  |
| Pericardial | 1 (5 %) | 0 |  | 0 |  |

4 ROIs in the left ventricle from short axis views were obtained for native T<sub>1</sub>, T<sub>2</sub>, and ECV. Healthy controls underwent CMR without contrast and therefore do not have ECV or LGE assessments. \*Insufficient T<sub>2</sub> mapping was performed on tactical athletes for comparison. ECV reference normal for healthy controls (<sup>b</sup>) and athletic controls (<sup>c</sup>) were derived from published literature, respectively.(9,10) <sup>¶</sup>Student t-test used for analysis.

Two COVID-19+ athletes had normal ECG, troponin I, and echocardiography with strain imaging (global longitudinal strain < −20 for both), but abnormal CMRs with inferoseptal LGE. One subject was diagnosed with acute pericarditis based on clinical symptoms (exertional chest tightness) after CMR demonstrated a pericardial effusion, pericardial LGE, and intramural LGE of the inferoseptum (**Figure 1**). The second subject was asymptomatic with undetectable troponin I, minor T wave abnormalities considered acceptable in trained athletes (and similar to priors), and normal echocardiogram with strain (LVEF > 65%, GLS-20.2). CMR showed mildly reduced biventricular systolic function (LVEF 50%, RVEF 46%) and acute myocarditis of the basal inferoseptum based on modified Lake Louise criteria.(15) While the septal and lateral wall ROIs in the patient with myocarditis were normal, ROIs in the area of LGE demonstrated elevations in all parameters (Native T_1_ 1184 ms, T_2_ 78.2 ms, ECV 39.2%; **Figure 1**). **Table 3** demonstrates comparison between COVID-19+ athletes with abnormal and normal CMRs.

**Table 3.**
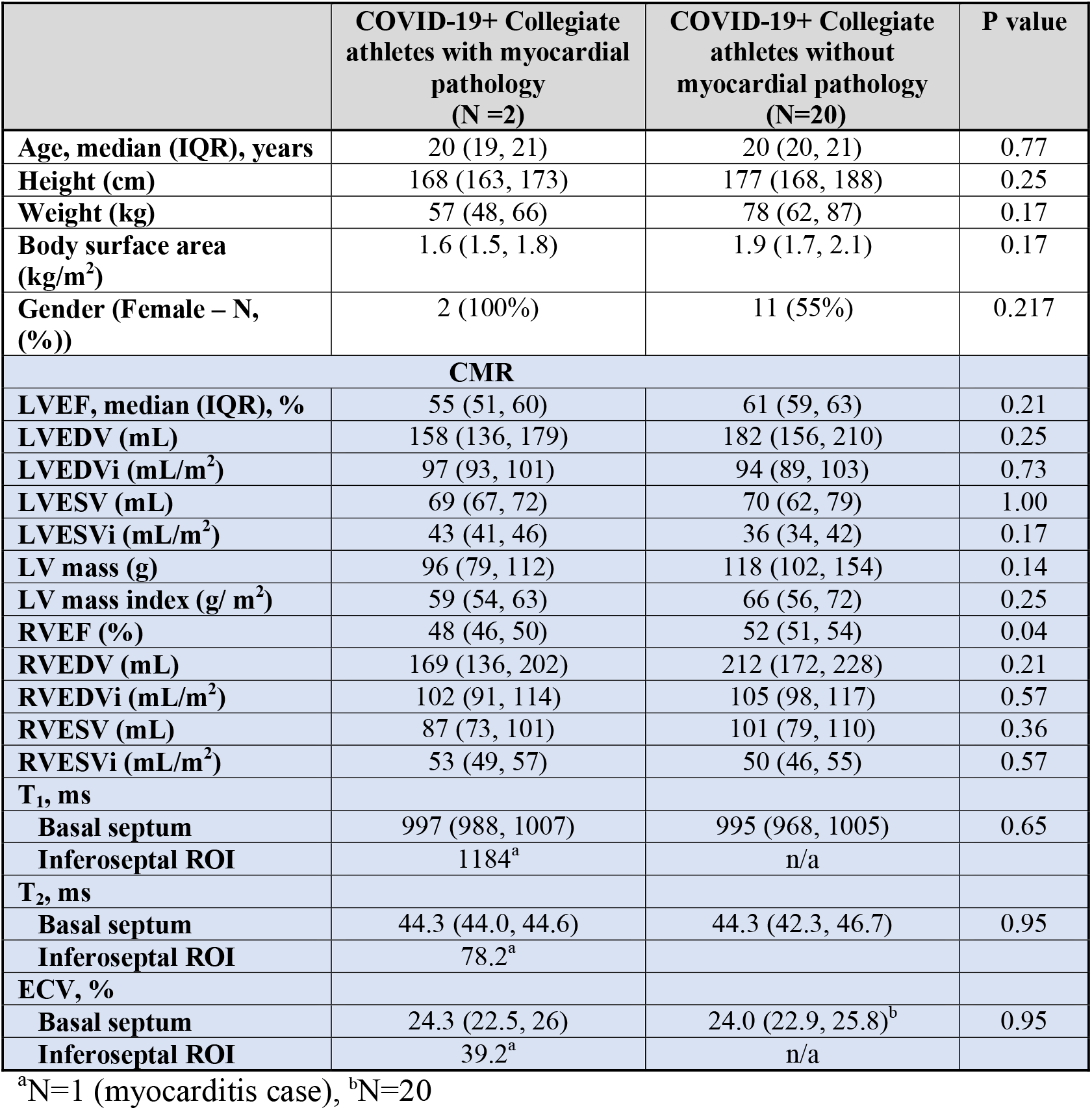
Comparison of COVID-19 positive collegiate athletes with and without cardiovascular involvement on CMR.

|  | **COVID-19+ Collegiate athletes with myocardial pathology (N =2)** | **COVID-19+ Collegiate athletes without myocardial pathology (N=20)** | **P value** |
| --- | --- | --- | --- |
| **Age, median (IQR), years** | 20 (19, 21) | 20 (20, 21) | 0.77 |
| **Height (cm)** | 168 (163, 173) | 177 (168, 188) | 0.25 |
| **Weight (kg)** | 57 (48, 66) | 78 (62, 87) | 0.17 |
| **Body surface area (kg/m<sup>2</sup>)** | 1.6 (1.5, 1.8) | 1.9 (1.7, 2.1) | 0.17 |
| **Gender (Female – N, (%))** | 2 (100%) | 11 (55%) | 0.217 |
| **CMR** |  |  |  |
| **LVEF, median (IQR), %** | 55 (51, 60) | 61 (59, 63) | 0.21 |
| **LVEDV (mL)** | 158 (136, 179) | 182 (156, 210) | 0.25 |
| **LVEDVi (mL/m<sup>2</sup>)** | 97 (93, 101) | 94 (89, 103) | 0.73 |
| **LVESV (mL)** | 69 (67, 72) | 70 (62, 79) | 1.00 |
| **LVESVi (mL/m<sup>2</sup>)** | 43 (41, 46) | 36 (34, 42) | 0.17 |
| **LV mass (g)** | 96 (79, 112) | 118 (102, 154) | 0.14 |
| **LV mass index (g/ m<sup>2</sup>)** | 59 (54, 63) | 66 (56, 72) | 0.25 |
| **RVEF (%)** | 48 (46, 50) | 52 (51, 54) | 0.04 |
| **RVEDV (mL)** | 169 (136, 202) | 212 (172, 228) | 0.21 |
| **RVEDVi (mL/m<sup>2</sup>)** | 102 (91, 114) | 105 (98, 117) | 0.57 |
| **RVESV (mL)** | 87 (73, 101) | 101 (79, 110) | 0.36 |
| **RVESVi (mL/m<sup>2</sup>)** | 53 (49, 57) | 50 (46, 55) | 0.57 |
| **T<sub>1</sub>, ms** |  |  |  |
| **Basal septum** | 997 (988, 1007) | 995 (968, 1005) | 0.65 |
| **Inferoseptal ROI** | 1184<sup>a</sup> | n/a |  |
| **T<sub>2</sub>, ms** |  |  |  |
| **Basal septum** | 44.3 (44.0, 44.6) | 44.3 (42.3, 46.7) | 0.95 |
| **Inferoseptal ROI** | 78.2<sup>a</sup> |  |  |
| **ECV, %** |  |  |  |
| **Basal septum** | 24.3 (22.5, 26) | 24.0 (22.9, 25.8)<sup>b</sup> | 0.95 |
| **Inferoseptal ROI** | 39.2<sup>a</sup> | n/a |  |

<sup>a</sup>N=1 (myocarditis case), <sup>b</sup>N=20

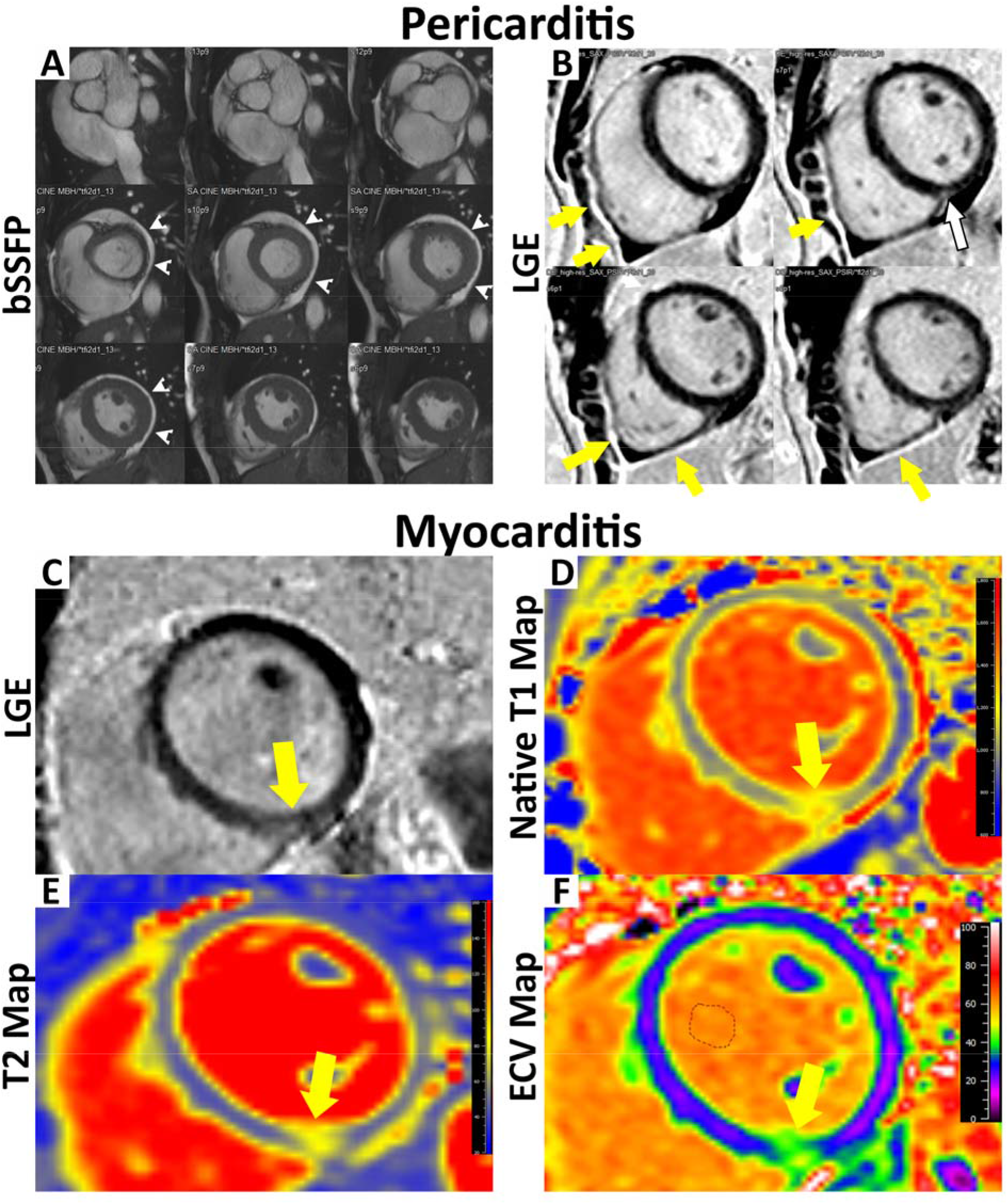
Central Illustration: Abnormal CMR in COVID-19+ athletes. A: Short axis stack with circumferential pericardial effusion (white arrowheads). B: Short axis stack late gadolinium enhancement phase-sensitive inversion recovery (PSIR) image with parietal pericardial LGE (yellow arrows) and basal inferoseptal LGE (white arrow). C: Short axis is PSIR with basal inferoseptal LGE (yellow arrow). D: Basal native T_1_, E: T_2_, and F: ECV maps with inferoseptal regional elevation in relaxation times. ROI in the areas of elevation demonstrated Native T_1_ 1184 ms, T_2_ 78.2 ms, ECV 39.2%.

Additionally, one COVID-19+ athlete had abnormal LVEF (<55%) and 4-chamber enlargement consistent with athlete’s heart. When compared with our laboratory’s normal values for parametric mapping, 10 COVID-19+ athletes (45%) with otherwise normal CMR exams had mild segmental elevations (**Table 4**), particularly of T_2_ times. Of note, three (13.6%) healthy controls and five (23%) tactical athletes also had mild segmental parametric mapping elevations.

**Table 4.**
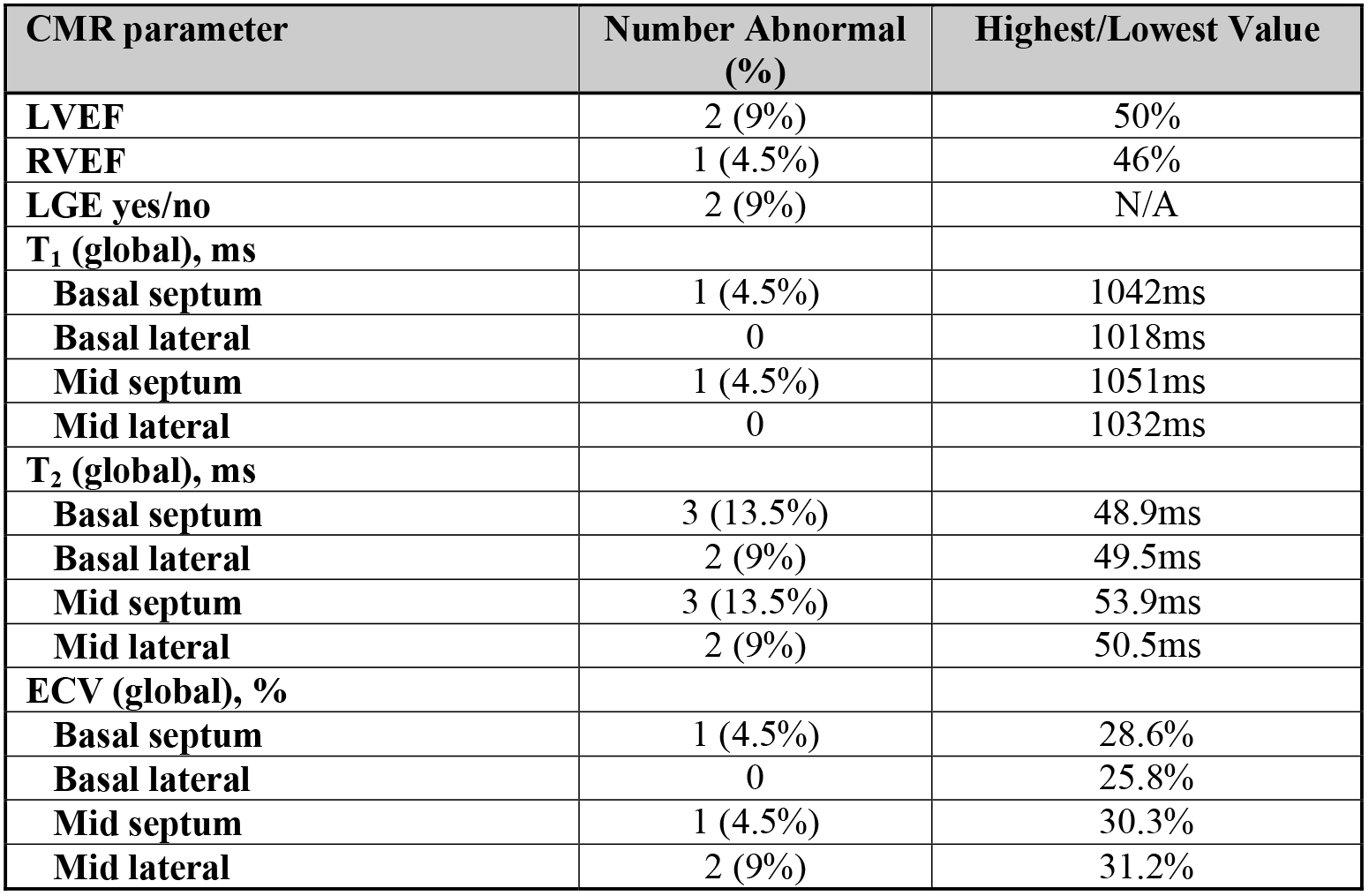
Abnormal CMR parameters among COVID-19+ athletes.

| **CMR parameter** | **Number Abnormal (%)** | **Highest/Lowest Value** |
| --- | --- | --- |
| **LVEF** | 2 (9%) | 50% |
| **RVEF** | 1 (4.5%) | 46% |
| **LGE yes/no** | 2 (9%) | N/A |
| **T<sub>1</sub> (global), ms** |  |  |
| **Basal septum** | 1 (4.5%) | 1042ms |
| **Basal lateral** | 0 | 1018ms |
| **Mid septum** | 1 (4.5%) | 1051ms |
| **Mid lateral** | 0 | 1032ms |
| **T<sub>2</sub> (global), ms** |  |  |
| **Basal septum** | 3 (13.5%) | 48.9ms |
| **Basal lateral** | 2 (9%) | 49.5ms |
| **Mid septum** | 3 (13.5%) | 53.9ms |
| **Mid lateral** | 2 (9%) | 50.5ms |
| **ECV (global), %** |  |  |
| **Basal septum** | 1 (4.5%) | 28.6% |
| **Basal lateral** | 0 | 25.8% |
| **Mid septum** | 1 (4.5%) | 30.3% |
| **Mid lateral** | 2 (9%) | 31.2% |

## DISCUSSION

There is a paucity of data regarding the cardiac sequelae of SARS-CoV-2 infection among athletes. This study of collegiate athletes demonstrated that the prevalence of myocardial inflammation or fibrosis after an asymptomatic or mild course of ambulatory COVID-19 is modest (9%). However, it also highlights that screening programs with less intensive cardiovascular phenotyping(16) are likely to underdiagnose myopericardial inflammation, as both COVID-19+ athletes with LGE on CMR had reassuring ECG, troponin I, and echocardiography with strain. In our cohort, using echocardiography cut-offs of LVEF < 55% and global longitudinal strain > −17% would have resulted in four false positives and a 0% positive predictive value for myopericarditis.

Many of the volumetric and parametric mapping changes seen between COVID-19+ athletes and healthy controls were less pronounced when compared to a group of tactical athletic controls (**Table 1**). T_2_ relaxation times (46.4 vs. 44.6 ms) were elevated among COVID-19+ athletes relative to healthy controls. No comparison could be made to tactical athletes, as they did not have clinical indication for T_2_ mapping. The limited literature on T_2_ mapping among athletes suggests values are slightly elevated compared to controls.(17,18) These findings highlight many of the known cardiac volumetric and functional changes associated with athleticism and emphasize the need for using athletic case-controls in future studies of COVID-19+ athletes.(11)

Forty-five percent of COVID-19+ athletes had mild segmental abnormalities on parametric mapping (**Table 4**). However, given the large number of segments evaluated, and similar abnormalities seen in the control groups, much of these differences may be explained by random chance. These abnormalities in isolation are of uncertain significance and in the authors’ opinion do not merit suspension of return to competition. However, even in the absence of symptoms of other screening abnormalities (ECG, troponin I, echocardiography with strain), 9% of COVID-19+ athletes had LGE, which has repeatedly been shown to predict mortality among patients with cardiovascular disease, including myocarditis, and should result in suspension of competition.(19,20) The ECV was found to be higher among COVID-19+ athletes (25.4%) compared to tactical athletic controls (22.5%) and healthy controls (24.0%), though the latter did not reach statistical significance. Due to myocyte hypertrophy, the ECV in athletes inversely correlates with the level of training.(10,11) Thus, this finding suggests a residual expansion of the extracellular space following COVID-19 infection. However, without pathological specimens, definitive analysis of these tissue-level changes cannot be definitively determined.

Cardiac involvement by COVID-19 is known to occur both in the acute and early convalescent phases and may result from direct viral-mediated cardiomyocyte injury, innate inflammatory reaction to infection, or a delayed, dysregulated immune response to COVID-19 (as seen in MIS-C).(21,22) Among symptomatic, hospitalized patients with COVID-19, CMR revealed myocardial edema in over half, and LGE in nearly one-third of all patients.(23) The largest cohort of COVID-19+ patients assessed by CMR consisted of 100 patients, one-third of whom required hospitalization, while the remainder recovered at home.(4) In that study, 78% of patients had an abnormal finding on CMR. The most common abnormality was persistent elevation of the native T_1_ and T_2_ relaxation times (occurring in 73% and 60% of patients, respectively).

There are many plausible explanations for the differences between the findings of that study and our cohort. First, our population was younger (median age 20 vs. 49 years old) and healthier with fewer cardiac risk factors at baseline. Second, there is a programmatic effort at our university for screening of student-athletes, and all that tested positive for COVID-19 are mandated to undergo CMR prior to returning to play, regardless of the severity of illness or symptoms. In the observational cohort in Germany, patients were referred for CMR on the clinical judgment of their physician, creating a selection bias. For example, their subjects were much more likely to have cardiac biomarker evidence of myocardial damage whereas no subjects in our cohort had elevated troponin. Thus, the German cohort represented a less healthy population who suffered a higher severity of illness. Another important difference between the two studies is that the median duration from illness to CMR in our study was only 52 days compared to 71 days in their study. This shorter duration between illness and CMR would be expected to increase the sensitivity of CMR for the detection of residual inflammation or fibrosis in our cohort. However, as noted above, our cohort had only a modest incidence of abnormalities on CMR.

It should be noted that this study began prior to the return of all students to campus. It is quite likely that cases will increase as students congregate again on campus, despite all reasonable measures to maintain social distancing. As new cases arise and screening continues, the duration between illness and CMR will naturally decrease, and thus the incidence of residual inflammation or LGE on CMR may increase. This may have a direct effect on athlete eligibility. Finally, all patients in the present study were asymptomatic or mildly symptomatic from their COVID-19 illness. Those with normal CMRs have been cleared for graded, progressive return to sport with ongoing monitoring for symptoms. The two athletes who tested positive are being withheld from training, as per the published guidelines. The degree of cardiac involvement from moderate-severe courses of COVID-19 among competitive athletes remains unknown.

### Limitations

There are inherent limitations of the retrospective design of this study. First, clinical outcomes among the collegiate athletes with and without CMR findings are not yet available. While COVID-19+ athletes were mandated to have laboratory, electrocardiographic, echocardiographic, and CMR imaging performed prior to return to sports participation, CMR was prioritized and some of this testing is incomplete at the current time. Baseline athletic performance phenotyping (quantification of baseline aerobic and strength training activities per week, peak VO2 with cardiopulmonary exercise testing, etc.) was not studied. While our cohorts’ diversity of sports and size limits our ability to determine if the type or aerobic intensity of sport affects the implications of the cardiovascular sequelae following COVID-19 infection, CMR volumetrics of our cohort point towards athlete’s heart (increased LV mass and increased four-chamber volumes), which as generalizability to many US competitive athletes. Our study did not enroll children < 18 years old, and therefore caution should be taken with generalization of these results to that age group.

## CONCLUSIONS

Our study suggests that the prevalence of myocardial inflammation or fibrosis after an asymptomatic or mild course of ambulatory COVID-19 among competitive athletes is modest (9%), but would be missed by ECG, Ti, and strain echocardiography. However, the burden of cardiovascular sequelae is still significant among this healthy cohort with minimal COVID-19 symptomatology, and the prevalence of myocardial involvement may be higher among those with more severe infections. Ongoing investigation is necessary to further phenotype the cardiovascular manifestations of COVID-19 in all phases of infection, and to study the potential clinical outcomes ranging from limitations in athletic performance to arrhythmia, heart failure, and sudden cardiac death in athletes and the general population. Additionally, the sensitivity of conventional screening with electrocardiography, cardiac biomarkers, and echocardiography needs to be compared to CMR in larger cohorts of competitive athletes to better understand the ideal screening mechanism to guide safe return to competition.

## PERSPECTIVES

### Competency in Patient care and medical knowledge

Cardiovascular sequelae of COVID-19 asymptomatic or mild illness among competitive athletes is lower than expected, but still nearly 10% and can present without associated symptoms.

### Translational Outlook

The sensitivity of current screening methods of electrocardiography, cardiac biomarkers, and echocardiography needs to be compared to CMR in larger cohorts of competitive athletes to better understand the ideal screening mechanism for return to competition.

## Data Availability

All data is stored in a de-identified database in REDCap.

## ACKNOWLEDGEMENTS

The authors would like to acknowledge our exceptional cardiac magnetic resonance technologists (Shannon Bozeman, Dana Fuhs, and Matt Herscher) for obtaining phenomenal imaging and working over-time for the screening of these athletes.

## Disclosures

All authors have no disclosures to report, including any relationship to industry.

**Abbreviations**
CMR: cardiac magnetic resonance
COVID-19: coronavirus disease-19
ECG: electrocardiogram
ECV: extracellular volume
LGE: late gadolinium enhancement
MIS-C: multisystem inflammatory syndrome
MOLLI: modified Look-Locker inversion recovery
ROI: regions of interest
SARS-CoV-2: severe acute respiratory syndrome coronavirus-2
SD: standard deviation

